# Iron biomarkers predict peripheral artery disease in females: a cross-sectional analysis of HEIST-DiC and NHANES participants

**DOI:** 10.1101/2023.08.21.23293418

**Authors:** Anand Ruban Agarvas, Stefan Kopf, Tiago J.S. Lopes, Paul Thalmann, José Manuel Fernández-Real, Peter Nawroth, Martina U. Muckenthaler

**Affiliations:** Heidelberg University, Heidelberg, Germany; Center For Translational Biomedical Iron Research, Department of Pediatric Hematology, Oncology Immunology and Pulmonology, University Hospital Heidelberg, Heidelberg, Germany; Clinic for Endocrinology, Diabetology, Metabolic Diseases and Clinical Chemistry (Internal Medicine 1), University Hospital Heidelberg, Heidelberg, Germany; German Center of Diabetes Research (DZD), Neuherberg, Germany; Institute of Medical Biometry, University Hospital Heidelberg, Heidelberg, Germany; Department of Diabetes, Endocrinology and Nutrition, Institut d’Investigació Biomèdica de Girona (IdIBGi), CIBEROBN (CB06/03/010) and Instituto de Salud Carlos III (ISCIII), Girona 17007, Spain; Molecular Medicine Partnership Unit, Heidelberg, Germany; Translational Lung Research Center Heidelberg (TLRC), German Center for Lung Research (DZL), Heidelberg, Germany; German Centre for Cardiovascular Research (DZHK), Partner Site Heidelberg/Mannheim, Germany; Nezu Life Sciences, Karlsruhe, Germany

**Keywords:** Iron, Vascular Diseases, Arterial Occlusive Diseases, Atherosclerosis, Peripheral Arterial Disease

## Abstract

**Background:** Iron overload has been proposed as a risk factor for atherosclerosis but the available data are controversial. Here we investigated whether iron biomarkers within physiological limits that frequently differ between sexes are associated with peripheral arterial disease (PAD)

**Methods:** Using two different analytical approaches (machine-learning and logistic regression), we studied the association between blood iron biomarkers and PAD in 368 individuals from the Heidelberg Study on Diabetes and Complications (HEIST-DiC) and in 5101 individuals from the National Health and Nutrition Examination Survey (NHANES 1999-2004).

**Results:** We found that iron biomarkers were among the top predictors of PAD in the machine-learning classification in both cohorts. In the HEIST-DiC cohort, ferritin, iron, and transferrin were ranked among the top predictive markers, while in the NHANES cohort ferritin, total iron binding capacity (TIBC), transferrin saturation (TSAT) and iron showed high predictive power. In the regression analysis ferritin showed a positive interaction among females in the HEIST-DiC (OR 2.68, 95% CI 0.94-7.61, *p=*0.057) and NHANES cohorts (OR 1.76, 95% CI 1.16-2.67, *p*=0.008). The multivariable regression analysis of the NHANES cohort, detected a nonlinear relationship between ferritin and PAD in that certain ferritin ranges (48-97 ng/mL: OR 14.59, 95% CI 1.6-135.93, p= 0.019; 98-169 ng/mL: OR 171.07, 95% CI 1.27-23404, p=0.039) in females associated positively with PAD.

**Conclusion:** Taken together our data show that iron biomarkers, importantly, elevated ferritin is associated with clinically apparent PAD in females. These findings add to the body of evidence suggesting sex differences in PAD and highlight a possible role of iron (directly or indirectly) in this relationship.

## Introduction

Peripheral arterial disease (PAD) is a common atherosclerotic vascular disorder of the arteries of the lower limbs that affects millions of individuals worldwide^1,2^. Compared to cardiovascular and cerebrovascular disease, PAD is less well studied, and our understanding of the disease process is still poor. The known risk factors of PAD include old age, smoking, and comorbidities such as diabetes, hypertension or Vitamin D excess^1^.

Iron is an essential micronutrient, however, its accumulation in excess contributes to the generation of reactive oxygen species. Various regulatory systems maintain iron homeostasis by controlling absorption, transport, and storage^3^. Under physiological conditions, iron is bound to transferrin in the plasma, and transported to cells that require iron. Usually, less than one-third of transferrin’s binding sites are occupied by iron, allowing free iron entering the circulation to be efficiently sequestered. However, in pathological states associated with iron overload, the binding capacity of transferrin is nearly saturated resulting in the appearance of ‘non-transferrin-bound iron (NTBI)’. Therefore, iron overload is discussed as a risk factor for the development of atherosclerosis^4^. Accordingly, inhibition of ferroptosis, an iron-dependent form of cell death, has been shown to protect from atherosclerosis^5^. Iron and PAD may be linked by sex hormones. Changes in estrogen and progesterone levels associated with menopause were shown to impair endothelial function, especially among women^6,7^. Both hormones further control hepcidin, the master regulator of iron homeostasis in opposing directions, with estrogen^8^ inhibiting and progesterone^9^ activating hepcidin expression.

In our previous work, we demonstrated that iron overload in mouse models and humans with hemochromatosis show an aggravation of atherosclerosis and vessel damage, respectively^10^. Specifically, we found that NTBI in the circulation contributes to atherosclerosis by oxidising low-density lipoproteins (LDL) and by inducing vascular dysfunction; in addition, iron deposition in the vasculature of hemochromatotic mice was associated with more severe atherosclerosis^10^. Whether or not iron levels within physiological limits affect atherogenesis is currently unclear and available data are controversial. Most studies undertaken so far have investigated the relationships between iron-related parameters [e.g. plasma iron, ferritin, transferrin, transferrin saturation (TSAT)] and cardiovascular disease^11–17^ but associations with PAD are rarely addressed.

To address this, we analyzed iron biomarkers for their associations with PAD and by considering sex-specific differences. We used two different datasets and orthogonal approaches: 1) machine learning (ML) classification and 2) logistic regression. Both methods can be used for analyzing relationships between variables, but they differ in their approach and application. ML classification tasks can analyze complex patterns in multidimensional data and identify nonlinear interactions among variables. They are particularly useful when manual specification of relationships is challenging as is the case for human clinical data. In contrast, regression analysis quantifies the strength and direction of associations between variables and provides interpretable results in terms of odds ratios. Both approaches carry their strengths and limitations. However, we expect that true associations in the data (in this case, sex-specific differential associations between iron biomarkers and PAD) should be detected by both methods. Therefore, we initially used the ML approach to provide unbiased insight into our data and subsequently regression analyses to validate identified relationship. An outline of the study is shown in Figure 1.

**Figure 1.**
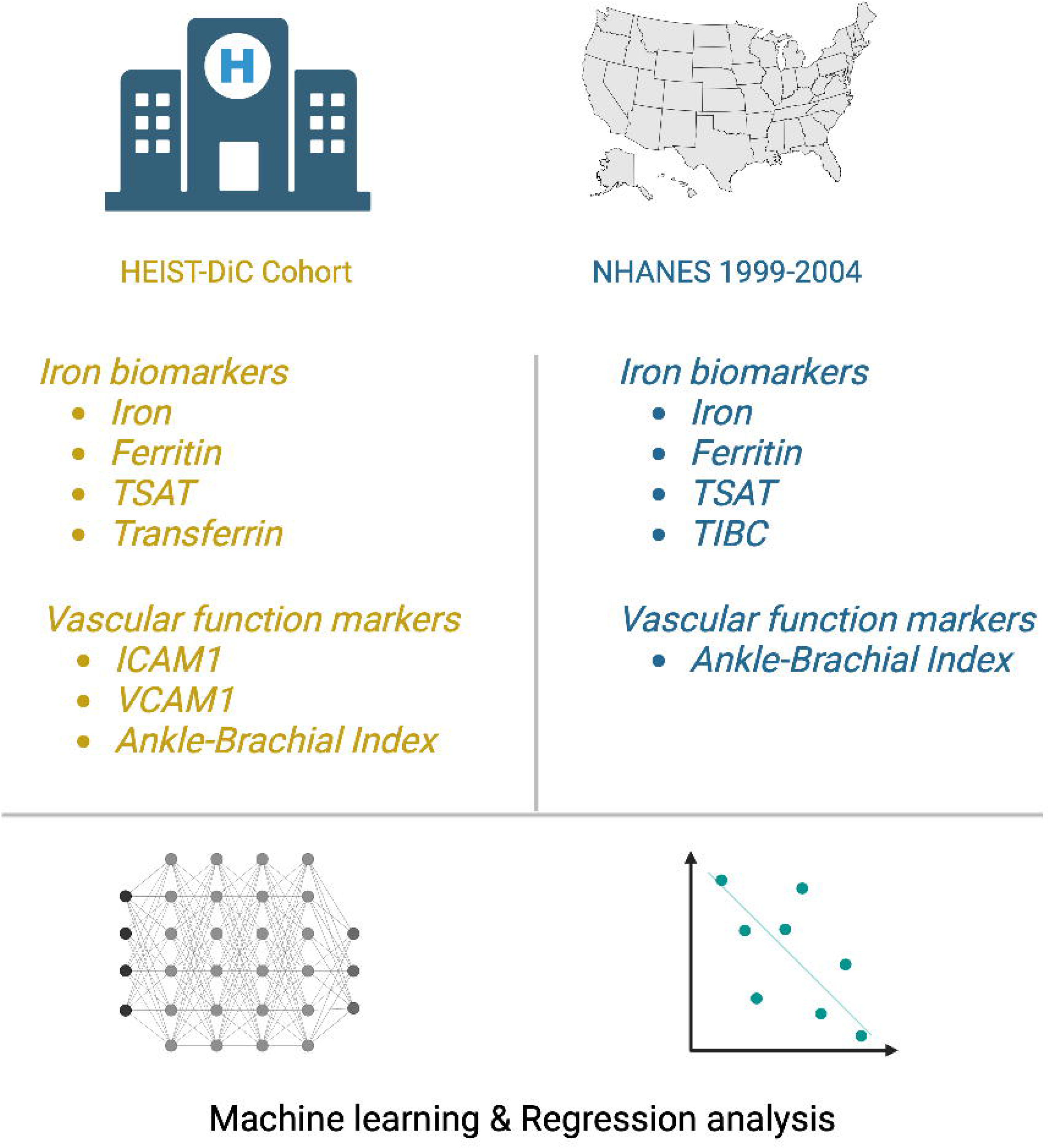
Outline of the study and analysis approach.

## Methods

### HEIST-DiC study

The Heidelberg Study on Diabetes and Complications (HEIST-DiC; https://clinicaltrials.gov; NCT03022721) is a prospective, observational study designed to analyze the development of complications in individuals with diabetes and prediabetes. We analyzed the data from 368 individuals enrolled in the HEIST-DiC study. The selection criteria for participants have been described previously^18^. We excluded participants with values of serum iron biomarkers suggestive of hemochromatosis^19^. In addition, since ferritin not only indicates iron stores, but also is an acute phase protein that responds to inflammation, we excluded participants with ferritin> 400 ng/mL. For all participants, a detailed medical history was obtained. The ankle-brachial index (ABI) was measured using noninvasive blood pressure measurements of arms and ankles (ABI System 1000; Boso d.o.o.). Blood samples were obtained in the fasting state and all baseline parameters were analyzed under standardized conditions in the central laboratory of the University Hospital of Heidelberg. Serum samples were stored at −80C until further analysis. We analyzed iron, ferritin, and TSAT as biomarkers of systemic iron status, and assayed serum intercellular adhesion molecule-1 (ICAM1) and vascular-cell adhesion molecule-1 (VCAM1) as markers of vascular dysfunction. We measured ICAM1 (#171B6009M) and VCAM1 (#171B6022M) in serum using multiplex magnetic bead-array-based technology on a BioPlex200 system (Bio-Rad) according to the manufacturer’s instructions. All participants provided written informed consent, and the ethics committees of the University of Heidelberg approved this study (Decision No. 204/2004, 400/2010, and S-383/2016) according to the Declaration of Helsinki.

### NHANES

The National Health and Nutrition Examination Survey (NHANES) is designed to assess the health and nutritional status of adults and children in the United States. The survey includes both an interview (in which participants are asked questions about their health and lifestyle), and a physical examination component (in which various measurements and samples are collected). We obtained data from 15,969 participants over 40 years in 3 consecutive cycles of NHANES (1999-2004) for whom ABI was measured.

The demographic variables, age (in years) and body mass index (BMI) were used as is. Smoking and alcohol were categorized according to a previous study^20^. We further categorized diabetes based on self-reported history, current use of insulin or oral hypoglycemic agents, fasting plasma glucose (FPG) ≥7.0 mM, or glycated hemoglobin (HbA1c) ≥6.5%. Prediabetes was defined among those with an absent self-reported history of diabetes by either FPG 5.6-6.9 mM or HbA1c 5.7-6.4%. We categorized hypertension based on self-reported history or current use of anti-hypertensive agents or systolic blood pressure >130 mmHg or diastolic blood pressure >80 mmHg. The presence of nonalcoholic fatty liver disease (NAFLD) was categorized by calculating the Fibrosis-4 Index score (FIB-4 score>3.25)^21^. As an index of insulin resistance, we calculated HOMA2-IR using C-peptide values instead of insulin^22^. The use of antiplatelet and antihyperlipidemic medications was categorized based on medication intake.

We applied the following exclusion criteria: not even one ABI measurement available, likely hemochromatosis^19^, anemia (hemoglobin <10g/L), elevated ferritin (>400 ng/mL), cancer, on current hormonal medications or iron supplements, donated blood in the last 1 month, with features of kidney disease (on dialysis or severe albuminuria), pregnant or breastfeeding in the last year, high risk of liver fibrosis, concurrent inflammation [C-Reactive Protein (CRP) >3 mg/dL] and with a feature of familial hyperlipidemia [LDL-Cholesterol (LDLc) >4.9 mM]. This resulted in a final sample number of 5101 participants. We obtained data from the following iron biomarkers: iron, ferritin, total iron binding capacity (TIBC), and TSAT. Assays for the analytes in NHANES were performed under strict laboratory protocols (for more details on data fields and procedure manuals, see Supplementary Table 1).

### Categorization as PAD

In both cohorts, ABI values were measured on both the right and left limbs. We categorized PAD when the ABI value on one of the sides was <0.9 or >1.4 as per recommendations^23^.

#### Implementation of the ML framework

First, we preprocessed the HEIST-DiC and the NHANES dataset and removed all features that contained more than 10% of missing values. Additionally, to made both cohorts comparable by only considering the cases that did not have missing values for ferritin, diabetes, CRP and high blood pressure in the NHANES cohort. For the HEIST-DiC dataset, we have used the leave-one-out split strategy, and for each fold, we filled in the missing values using the average of each column (SimpleImputer method, from Scikit-learn). Moreover, we used SMOTE (v0.12.0)^24^ to oversample the minority class (PAD), to match the majority class (No PAD). For the NHANES dataset, we used the 10-fold cross-validation strategy, and for each fold, we filled the missing values and oversampled the minority class using the same methods, and performed the so-called one-hot encoding, where we encoded the categorical variables into numerical values. Next, for each dataset separately, we trained the XGBoost classifier (v2.0.3)^25^ and predicted the class of the instances on the test set, from where we derived the balanced accuracy, the F1 Score, and quantified the feature importance. Finally, we also implemented a version of the ML framework that allowed us to assess the variable importance using SHAP (SHapley Additive exPlanations, v0.44.0).^26^ Here, we performed the same steps for both HEIST and NHANES datasets, except that we did not use any split strategy; instead, we filled the missing values, oversampled the minority class and trained the classifier using the complete dataset. The ML pipeline is shown in Figure 2A.

**Figure 2.**
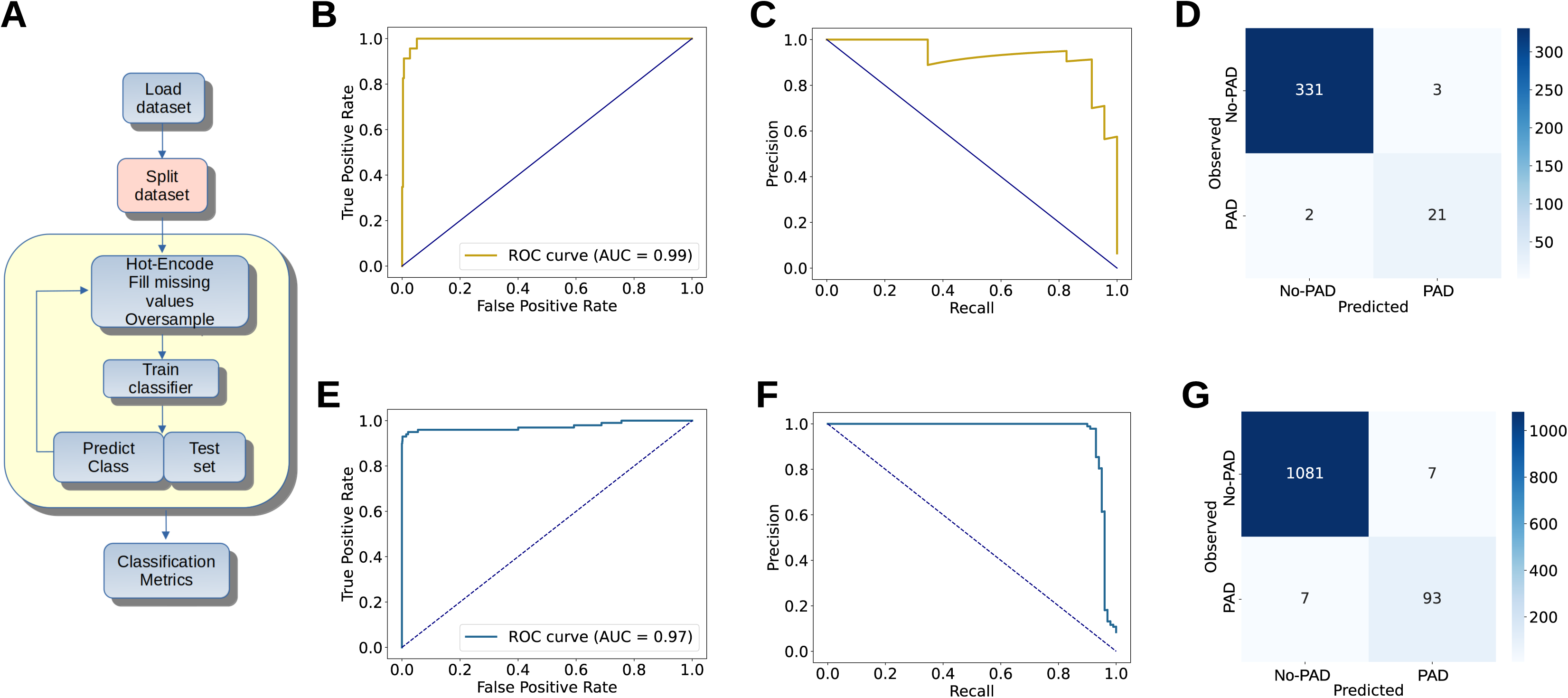
Setup and results of the machine learning approach. (A) The datasets were divided using either cross-fold validation (NHANES), or the leave-one-out strategy (HEIST-DIC). Next, the categorical variables were transformed into binary values (the so-called ‘hot-encode’), we filled missing values and oversampled the minority classes (see Methods). After training the classifier, the instances of the test set had their classes predicted, and we evaluated the results using classical ML metrics. (B-D) The ROC and precision-recall curves of the HEIST-DIC cohort data, and its confusion matrix. (E-G) The ROC and precision-recall curves of the NHANES cohort data, and its confusion matrix.

#### Regression analysis

For the regression analysis, we first tested the interaction effect of each of the iron biomarkers with sex in both cohorts as iron metabolism is hallmarked by sex-specific differences. Further, in the NHANES cohort, we conducted multivariable regression analyses by flexibly modelling the probability of PAD’s presence with one of the iron biomarkers (iron, ferritin, or TSAT) by spline regression (degree of freedom=4). We chose variables from the feature importance in ML analysis as covariates in the regression models on the NHANES data: age, CRP, HDLc, triacylglycerols, HbA1c, ACR, BMI, ethnicity (non-Hispanic whites as the reference), smoking (non-smokers as reference), the presence of diabetes, AST, ALT, Hb, creatinine, antihyperlipidemic medications; the former seven variables as continuous and the rest as categorical). All continuous variables were mean-centered for the analysis, and variables were included in the models only when their missing data was <10%. In addition, sample weights were included in the analyses of NHANES data.

All regression analyses were performed using R Statistical Software (v4.2.2^27^). We used Python (v3.10.12^28^), and Scikit-learn (v1.4.0^29^) for the implementation of the ML framework. The reporting of the study is as per The Strengthening the Reporting of Observational Studies in Epidemiology (STROBE) guidelines^30^. We have also developed a data exploration tool for readers to explore the datasets and interactively create their visualizations from the paper (https://anandr.shinyapps.io/NHANES_HEISTDiC/) using Quarto^31^ and Shiny^32^.

## Results

### Demographics

The baseline characteristics of the HEIST-DiC participants are shown in Table 1. The overall prevalence of PAD was 6.6% with a higher proportion among males (17/192) than females (6/165). Among both sexes, the participants with PAD were older and diabetes and hypertension was a common feature (Table 1). The sICAM1 and sVCAM1 levels were significantly higher only among males with PAD (Table 1). Among the iron biomarkers, ferritin was elevated among females, but not males, with PAD (Median 141, IQR 93-254, *p=0.035*) while iron, transferrin, and TSAT were similar between HEIST-DIC participants with and without PAD. (Table 1).

**Table 1.**
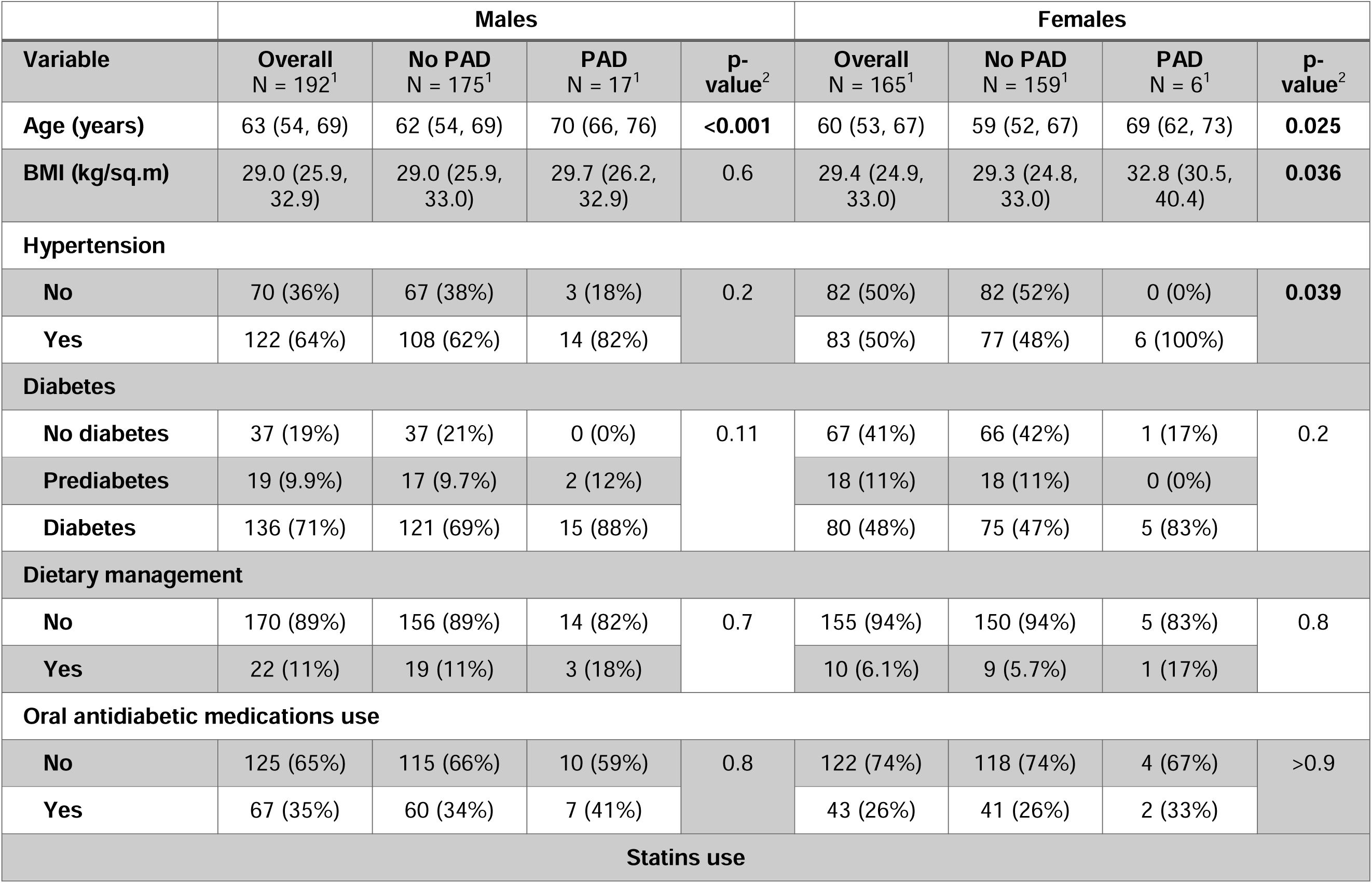
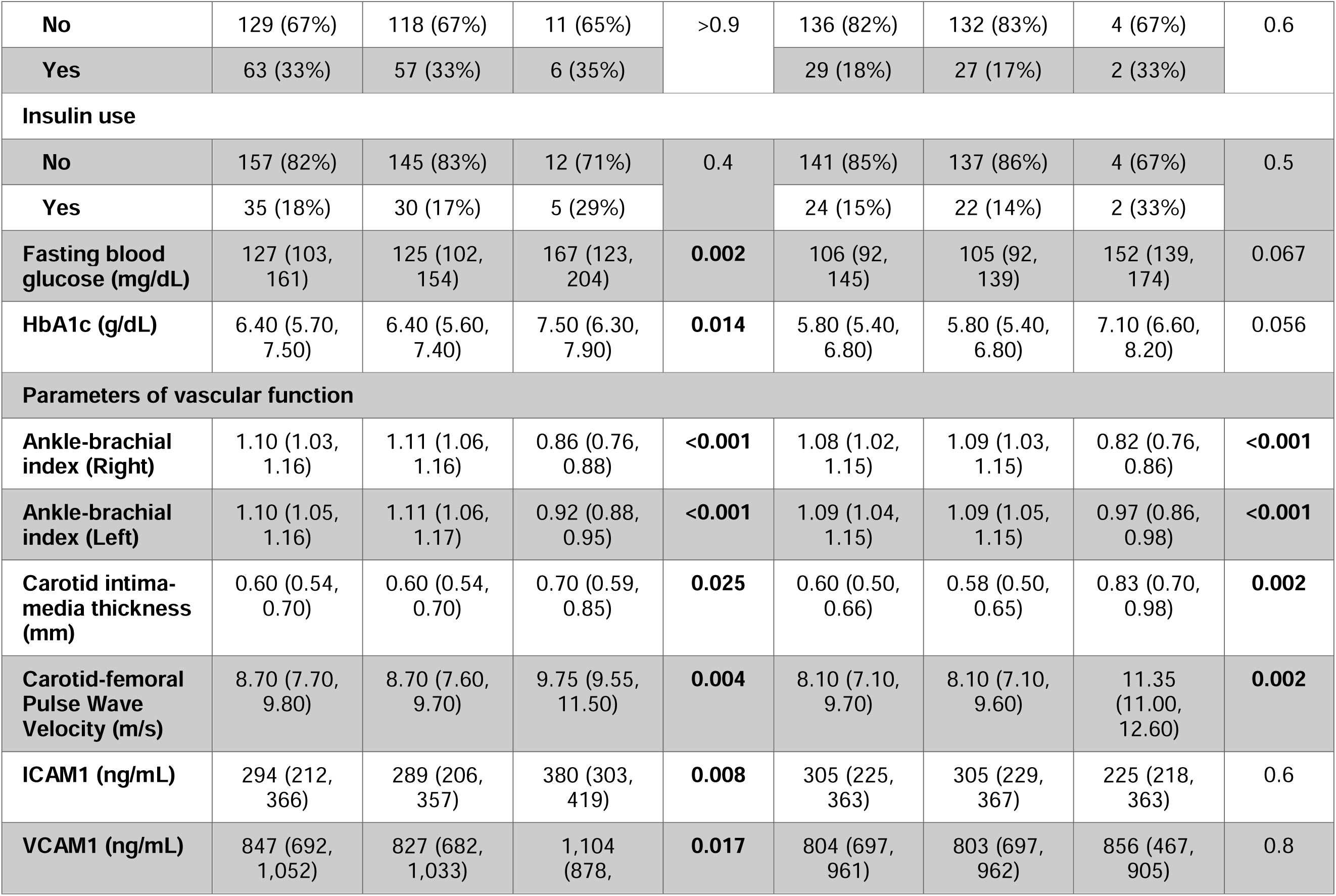
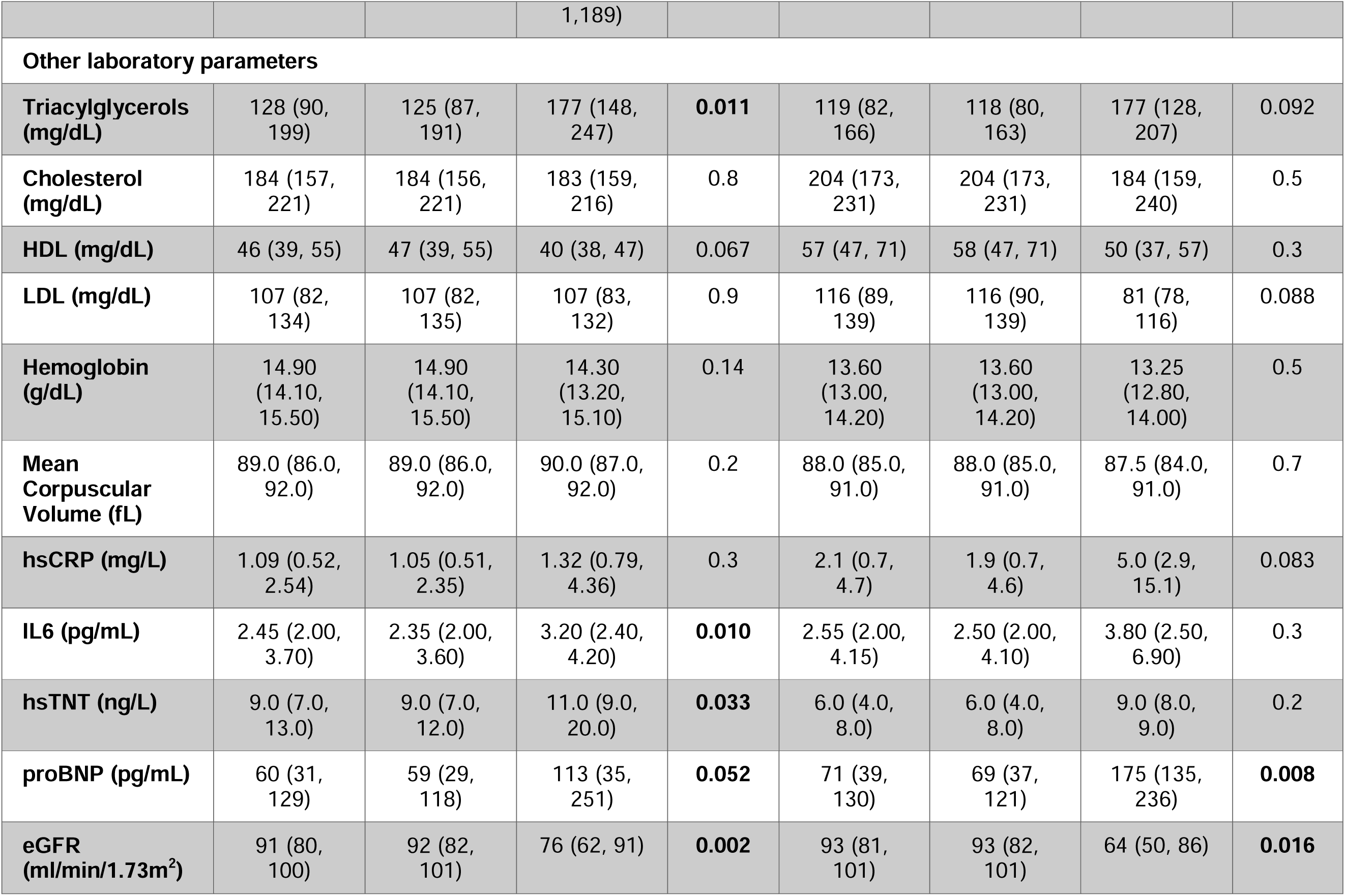
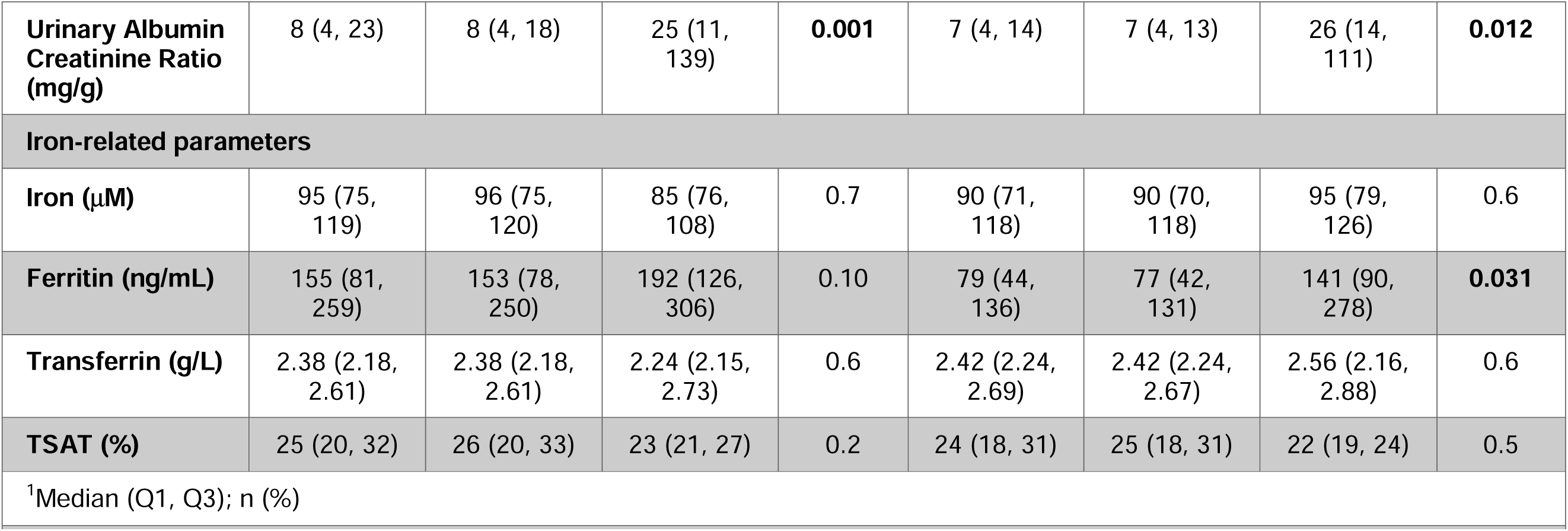
Demographics of HEIST-DiC participants. The demographics of the HEIST-DiC participants classified by sex and the presence of PAD are shown in Table 1. For continuous variables, the summary data are shown as Mean±Standard Deviation and for categorical variables as N (%). P-values as calculated by the Wilcoxon rank sum test or Pearson’s Chi-squared test^C^ and significant P-values are boldfaced. BMI: Body Mass Index, HDL: High-Density Lipoprotein, LDL: Low-Density Lipoprotein, HbA1c: Glycated hemoglobin, ICAM1: Intercellular Adhesion Molecule 1, VCAM1: Vascular Cell Adhesion Molecule 1, hsCRP: High sensitivity C-reactive protein, IL6: Interleukin 6, hsTNT: High sensitivity Troponin T, proBNP: N-Terminal B-type Natriuretic Peptide, eGFR: Estimated Glomerular Filtration Rate, TSAT: Transferrin Saturation

The analysis of NHANES builds on a previous report^33^ by including data from the NHANES 2003-2004 cycle. The baseline characteristics of the NHANES participants included in the study are shown in Table 2. Overall, the prevalence of PAD in the population was 6.25% with a significantly higher prevalence among females (males: 4.96%, females: 7.9%; *p*=0.001). Individuals with PAD showed differences in ethnicity, sex, presence of diabetes, smoking, alcohol status, physical activity, and use of antihyperlipidemic and antiplatelet medications (Table 2). Among the iron biomarkers, serum iron (Median 80, IQR 64-108 μg/dL; *p*<0.001) and transferrin saturation (Median 24, IQR 17-30; *p*=0.021) were reduced in males with PAD while females with PAD showed higher ferritin (Median 87, IQR 56-252 ng/mL; *p*<0.001) and lower TIBC (Median 60, IQR 54-68; *p*<0.001). Differences were also observed in some laboratory parameters within both sexes with PAD (e.g. albumin, AST, urea, creatinine, CRP, and ACR; Table 2).

**Table 2.** Demographics of NHANES 1999-2004 participants. The demographics of the NHANES 1999-2004 participants classified by sex and the presence of PAD are shown in Table 2. The ^1^N of participants shown are unweighted numbers. For continuous variables, the summary data are shown as Median (Interquartile Range), and for categorical variables as N (%)-survey weights were included in calculating the summary data. P-values as calculated by the Wilcoxon rank-sum test for complex survey samples or chi-squared test with Rao & Scott’s second-order correction^C^ and significant P-values are boldfaced. BMI: Body Mass Index, HDL: High-Density Lipoprotein, LDL: Low-Density Lipoprotein, HbA1c: Glycated hemoglobin, CRP: High sensitivity C-reactive protein, eGFR: Estimated Glomerular Filtration Rate, TSAT: Transferrin Saturation, TIBC: Total Iron Binding Capacity, AST: Aspartate Aminotransferase, ALT: Alanine Aminotransferase, FIB4 Score: Fibrosis-4 index Score

|  | Males |  |  |  | Females |  |  |  |
| --- | --- | --- | --- | --- | --- | --- | --- | --- |
| Characteristic | Overall<br>N = 2792<sup>1</sup> | No PAD, N<br>= 2573<sup>1</sup> | 1, N = 219<sup>1</sup> | p-<br>value<sup>2</sup> | Overall<br>N =2309<sup>1</sup> | 0, N =<br>2082<sup>1</sup> | 1, N = 227<sup>1</sup> | p-<br>value<sup>2</sup> |
| Ethnicity |  |  |  |  |  |  |  |  |
| Non-Hispanic<br>White | 1459<br>(78%) | 1330<br>(78%) | 129 (83%) | 0.003 | 1089<br>(73%) | 971 (72%) | 118 (76%) | <0.001 |
| Non-Hispanic<br>Black | 497 (9.0%) | 454 (8.9%) | 43 (11%) |  | 475 (11%) | 418 (11%) | 57 (16%) |  |
| Mexican-American | 662 (5.6%) | 619 (5.6%) | 43 (5.2%) |  | 546 (5.0%) | 504 (5.1%) | 42 (4.2%) |  |
| Others | 174 (7.6%) | 170 (7.9%) | 4 (1.2%) |  | 199 (11%) | 189 (12%) | 10 (4.0%) |  |
| Age (years) | 52 (45, 61) | 51 (45, 60) | 68 (60, 76) | <0.001 | 52 (45, 65) | 51 (45, 64) | 70 (54, 81) | <0.001 |
| BMI (kg/sq.m) | 27.9 (25.1,<br>31.0) | 27.9 (25.1,<br>30.9) | 27.8 (25.0,<br>32.2) | >0.9 | 27 (24, 32) | 27 (24, 32) | 29 (25, 34) | 0.034 |
| Diabetes (coded) |  |  |  |  |  |  |  |  |
| No diabetes | 1796<br>(75%) | 1698<br>(76%) | 98 (49%) | <0.001 | 1461<br>(73%) | 1358<br>(75%) | 103 (51%) | <0.001 |
| Prediabetes | 444 (13%) | 387 (12%) | 57 (27%) |  | 418 (16%) | 357 (15%) | 61 (25%) |  |
| Diabetes | 450 (12%) | 395 (12%) | 55 (24%) |  | 349 (11%) | 296 (10%) | 53 (23%) |  |
| Hypertension (coded) |  |  |  |  |  |  |  |  |
| No | 252 (14%) | 233 (14%) | 19 (13%) | 0.9 | 209 (15%) | 188 (15%) | 21 (14%) | 0.9 |
| Yes | 1557 | 1404 | 153 (87%) |  | 1296 | 1129 | 167 (86%) |  |
|  | (86%) | (86%) |  |  | (85%) | (85%) |  |  |
| Smoking (coded) |  |  |  |  |  |  |  |  |
| Nonsmoker | 974 (36%) | 931 (37%) | 43 (18%) | <0.001 | 1397 (58%) | 1277 (58%) | 120 (51%) | 0.089 |
| Former smoker | 1134 (39%) | 1023 (38%) | 111 (49%) |  | 532 (24%) | 466 (23%) | 66 (28%) |  |
| Current smoker | 680 (25%) | 615 (25%) | 65 (33%) |  | 375 (18%) | 335 (18%) | 40 (21%) |  |
| Alcohol drinker (coded) |  |  |  |  |  |  |  |  |
| No | 188 (33%) | 174 (33%) | 14 (39%) | 0.5 | 552 (44%) | 484 (43%) | 68 (50%) | 0.3 |
| Yes | 339 (67%) | 317 (67%) | 22 (61%) |  | 570 (56%) | 506 (57%) | 64 (50%) |  |
| Antihyperlipidemic medications (coded) |  |  |  |  |  |  |  |  |
| No | 2269 (91%) | 2083 (91%) | 186 (91%) | 0.9 | 1984 (95%) | 1788 (95%) | 196 (91%) | 0.037 |
| Yes | 177 (8.7%) | 159 (8.7%) | 18 (9.2%) |  | 113 (5.2%) | 97 (4.8%) | 16 (9.3%) |  |
| Antiplatelet medications (coded) |  |  |  |  |  |  |  |  |
| No | 2404 (99%) | 2209 (99%) | 195 (97%) | 0.006 | 2075 (99%) | 1867 (99%) | 208 (99%) | 0.3 |
| Yes | 42 (1.1%) | 33 (1.0%) | 9 (2.9%) |  | 22 (0.8%) | 18 (0.7%) | 4 (1.5%) |  |
| Ankle-brachial Index (Left) | 1.16 (1.09, 1.22) | 1.16 (1.10, 1.22) | 0.88 (0.79, 1.01) | <0.001 | 1.11 (1.04, 1.18) | 1.12 (1.06, 1.19) | 0.88 (0.78, 0.94) | <0.001 |
| Ankle-brachial | 1.16 (1.10, | 1.16 (1.11, | 0.87 (0.75, | <0.001 | 1.10 (1.03, | 1.11 (1.05, | 0.86 (0.77, | <0.001 |
| **Index (Right)** | 1.22) | 1.22) | 0.97) |  | 1.18) | 1.18) | 0.90) |  |
| **Albumin (g/L)** | 44.00<br>(42.00,<br>46.00) | 44.00<br>(42.00,<br>46.00) | 43.00<br>(41.00,<br>44.00) | **<0.001** | 43.0 (41.0,<br>44.0) | 43.0 (41.0,<br>45.0) | 42.0 (39.0,<br>44.0) | **0.001** |
| **AST (U/L)** | 26 (21, 33) | 26 (21, 34) | 22 (18, 27) | **<0.001** | 19 (15, 25) | 19 (15, 25) | 18 (16, 23) | 0.2 |
| **ALT (U/L)** | 24 (21, 29) | 24 (21, 29) | 23 (20, 28) | 0.059 | 21 (18, 25) | 21 (18, 25) | 22 (19, 26) | 0.15 |
| **Blood urea**<br>**nitrogen**<br>**(mmol/L)** | 5.00 (4.28,<br>6.10) | 5.00 (3.93,<br>6.07) | 6.07 (4.64,<br>7.14) | **<0.001** | 4.60 (3.57,<br>5.71) | 4.60 (3.57,<br>5.71) | 5.00 (3.90,<br>6.78) | **0.002** |
| **Creatinine**<br>**(μmol/L)** | 88 (71, 97) | 80 (71, 97) | 88 (80,<br>106) | **0.018** | 62 (53, 71) | 62 (53, 71) | 71 (62, 88) | **<0.001** |
| **Total**<br>**Cholesterol**<br>**(mg/dL)** | 203 (179,<br>228) | 204 (179,<br>228) | 193 (171,<br>220) | **0.008** | 207 (184,<br>235) | 207 (183,<br>235) | 211 (188,<br>238) | 0.3 |
| **Triacylglycerols**<br>**(mg/dL)** | 132 (89,<br>200) | 132 (89,<br>200) | 130 (86,<br>190) | >0.9 | 108 (73,<br>160) | 107 (71,<br>156) | 132 (95,<br>195) | **<0.001** |
| **LDL (mg/dL)** | 124 (103,<br>147) | 124 (103,<br>148) | 115 (85,<br>136) | **0.038** | 122 (101,<br>145) | 122 (101,<br>145) | 119 (97,<br>143) | 0.8 |
| **HDL (mg/dL)** | 44 (37, 54) | 44 (37, 54) | 44 (37, 52) | 0.4 | 55 (46, 67) | 56 (46, 67) | 52 (44, 61) | **0.006** |
| **Fasting plasma**<br>**glucose (mg/dL)** | 101 (94,<br>111) | 100 (94,<br>110) | 108 (92,<br>125) | 0.3 | 95 (90,<br>104) | 95 (90,<br>103) | 102 (96,<br>115) | **<0.001** |
| **Insulin (pmol/L)** | 57 (37, 92) | 56 (37, 90) | 85 (41,<br>129) | 0.089 | 48 (33, 80) | 47 (32, 77) | 99 (46,<br>130) | **0.007** |
| **CRP (mg/dL)** | 0.18 (0.08,<br>0.37) | 0.18 (0.08,<br>0.37) | 0.27 (0.14,<br>0.44) | **<0.001** | 0.25 (0.10,<br>0.58) | 0.25 (0.10,<br>0.57) | 0.32 (0.15,<br>0.76) | **0.036** |
| **Hemoglobin (g/dL)** | 15.30 (14.70, 16.00) | 15.30 (14.70, 16.00) | 15.00 (14.10, 15.80) | **0.006** | 13.90 (13.10, 14.50) | 13.90 (13.10, 14.50) | 13.80 (12.90, 14.70) | >0.9 |
| **FIB4 Score** | 1.14 (0.89, 1.48) | 1.13 (0.89, 1.47) | 1.28 (1.02, 1.62) | **0.002** | 0.85 (0.63, 1.15) | 0.84 (0.62, 1.14) | 1.07 (0.80, 1.37) | **<0.001** |
| **Albumin-creatinine ratio (μg/mg)** | 0.59 (0.40, 1.06) | 0.58 (0.40, 1.01) | 1.07 (0.63, 3.09) | **<0.001** | 0.81 (0.54, 1.54) | 0.79 (0.54, 1.44) | 1.30 (0.70, 2.67) | **<0.001** |
| **Iron (μM)** | 16.5 (12.9, 20.8) | 16.7 (12.9, 20.9) | 15.4 (11.6, 20.4) | 0.065 | 14.1 (10.6, 18.1) | 14.3 (10.6, 18.1) | 12.9 (10.2, 16.5) | **0.025** |
| **Ferritin (ng/mL)** | 142 (93, 223) | 142 (93, 223) | 120 (81, 197) | 0.3 | 60 (31, 109) | 60 (31, 107) | 85 (56, 192) | **<0.001** |
| **TSAT (%)** | 26 (20, 33) | 27 (20, 33) | 25 (17, 32) | 0.079 | 22 (16, 29) | 22 (16, 29) | 20 (16, 28) | 0.2 |
| **TIBC (μM)** | 63 (57, 70) | 63 (57, 69) | 63 (57, 70) | 0.8 | 64 (58, 71) | 64 (58, 71) | 61 (54, 70) | **0.001** |

#### Establishing a machine learning predictive framework for PAD

We next aimed to establish a predictive ML framework to anticipate the occurrence of PAD based on biomarkers. A so-called ML classifier is trained on a dataset containing instances with known outcomes (i.e., the presence or absence of PAD), and its relationship with input features. During the test phase, the trained classifier is used to predict PAD in participants unseen during the training phase, where we purposefully hid the true diagnostic. The result is then compared to the real diagnosis, and the quality of the framework is evaluated. Overall, this process enables the identification of subtle correlations between variables that might be imperceptible to human analysis, but that can be used to anticipate PAD and enhance our diagnostic capabilities.

Our framework received the available molecular and physiological biomarkers from participants from two distinct cohorts, HEIST-DiC and NHANES as input. These cohorts have unique characteristics that require the development of separate data preprocessing, training, and testing regimens. First, the HEIST-DiC cohort comprised 357 instances (23 PAD and 334 No PAD cases) and had 35 numerical and categorical features. Here, for the training and testing of the ML framework, we used a leave-one-out split methodology. This approach involves using each instance as a test case once, while the remainder forms the training set, thus maximizing the use of available data for training. In contrast, the NHANES cohort was considerably larger, with 1188 instances (100 PAD and 1088 No PAD), and 28 features; hence, we used the 10-fold cross-validation strategy. This method divides the dataset into ten parts, with each part successively used for testing while the other 9 parts are used for training. Together, these strategies ensured robustness and reliability in our model’s predictive performance.

We found that the ML framework achieved a balanced accuracy of 95% for the HEIST cohort (Figure 2B-D) and 96% for the NHANES cohort (Figure 2E-G). Balanced accuracy is an important metric in the context of imbalanced datasets, where one of the classes has many times more instances than the other (in the case of HEIST-DiC, No PAD cases were 14 times more frequent, and in NHANES, 10 times). Thus, the balanced accuracy represents the average accuracy obtained on both classes (PAD and No PAD), which in turn ensures that our model is equally adept at identifying both conditions.

Moreover, the ML framework also achieved high F1 scores of 0.89 for HEIST-DiC and 0.93 for NHANES, pointing to a strong balance between precision (the proportion of true positive results in all positive predictions), and recall (the proportion of true positive results in all actual positives). Thus, the F1 score in combination with the balanced accuracy provides realistic measures of a model’s strength, particularly in datasets where one class might significantly outnumber the other. In summary, the ML framework was effective in predicting the occurrence of PAD across different cohorts, even in the face of data imbalance. Further, sex and iron biomarkers were ranked among the top predictive markers for the presence of PAD in the HEIST-DiC (ferritin, iron and transferrin) and the NHANES cohorts (ferritin, TIBC, TSAT and iron; Supplementary Figure 2; Figure 3A-B).

**Figure 3.**
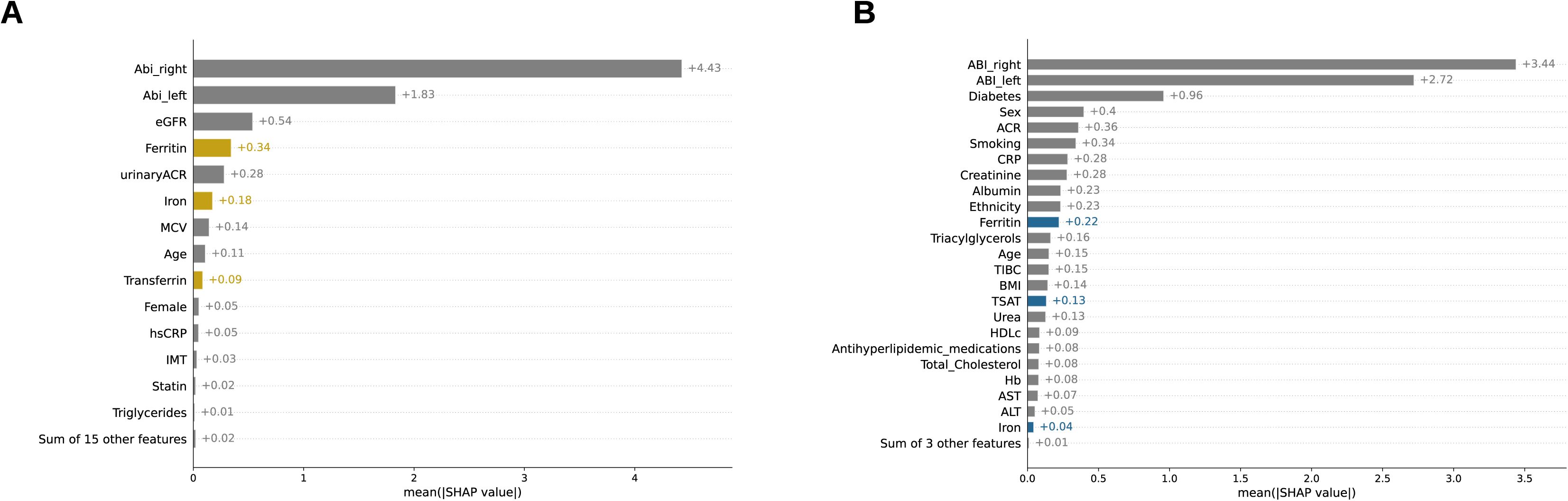
Feature importance of machine learning classification. (A-B) We used the SHAP method to quantify the importance of the features that were contributing the most to the prediction of PAD occurrence. Here, higher values indicate a higher contribution of a given feature. In both panels, iron-related markers are color-coded, and appear among the top features in terms of relevance to the prediction of PAD.

#### Regression analyses

We next performed regression analyses on the HEIST-DiC data and tested the interaction effect of each of the iron biomarkers with sex and found that ferritin showed a positive association in females with PAD (Ferritin*Sex, OR 2.68, 95% CI 0.94-7.61, *p=*0.057) while, iron (Iron*Sex, OR 1.17, 95% CI 0.44-2.96, p=0.7), transferrin (Transferrin*Sex, OR 1.28, 95% CI 0.48-3.17, *p=*0.6) and TSAT (TSAT*Sex, OR 1.1, 95% CI 0.35-3.2, *p=*0.9) were non-significant. Consistent with our observations in the HEIST-DiC cohort, we found that ferritin also showed a significant interaction with sex in the NHANES cohort (Ferritin*Sex, OR 1.76, 95% CI 1.16-2.67, *p*=0.008). TIBC also had a significant negative interaction (TIBC*Sex, OR 0.61, 95% CI 0.4-0.92, *p*=0.018), while iron (Iron*Sex, OR 0.95, 95% CI 0.69-1.31, *p*=0.76) and TSAT (TSAT*Sex, OR 1.15, 95% CI 0.84-1.58, *p*=0.37) were nonsignificant.

In the multivariable regression analyses of the NHANES data, we found that, ferritin or sex alone do not show a statistically significant relationship with PAD. On the other hand, the interaction between ferritin and sex reveals a significant positive association for two spline components (Table 3). Specifically, ferritin showed positive and statistically significant effects in females with two positions (48-97 ng/mL: OR 14.59, 95% CI 1.6-135.93, p= 0.019; 98-169 ng/mL: OR 171.07, 95% CI 1.27-23404, p= 0.039); this suggests that the relationship between PAD and ferritin is nonlinear and that within high ferritin ranges females show a positive association with PAD. Other significant covariates include age, CRP, and smoking, all of which also positively impact the outcome. By contrast, no associations were significant for iron and TSAT (Supplementary Table 2).

**Table 3.** Multivariable regression analysis of ferritin and PAD in the NHANES Cohort. We conducted multivariable regression analyses by flexibly modelling the probability of PAD’s presence with ferritin by spline regression (degree of freedom=4). The following variables were included a covariates: age, CRP, HDLc, triacylglycerols, HbA1c, ACR, BMI, ethnicity (non-Hispanic whites as the reference), smoking (non-smokers as reference), the presence of diabetes, AST, ALT, Hb, creatinine, antihyperlipidemic medications; the former seven variables as continuous and the rest as categorical). All continuous variables were mean-centered for the analysis. In addition, sample weights were included in the analyses BMI: Body Mass Index, HDL: High-Density Lipoprotein, CRP: High sensitivity C-reactive protein, AST: Aspartate Aminotransferase, ALT: Alanine Aminotransferase; ACR: Albumin-creatinine ratio, OR: Odds Ratio, 95% CI: 95% Confidence Interval, SE: Standard Error

| Table 3 | OR | SE | Z score | P. Value | 95% CI |  |
| --- | --- | --- | --- | --- | --- | --- |
|  |  |  |  |  | Low | High |
| Intercept | 0.001 | 3.205 | -2.338 | **0.019** | 0.000 | 0.297 |
| Ferritin (<48 ng/mL) | 0.868 | 0.896 | -0.158 | 0.875 | 0.150 | 5.031 |
| Ferritin (48-96 ng/mL) | 0.481 | 1.478 | -0.495 | 0.621 | 0.027 | 8.714 |
| Ferritin (97-168 ng/mL) | 0.284 | 2.034 | -0.619 | 0.536 | 0.005 | 15.306 |
| Ferritin (>169 ng/mL) | 0.777 | 1.061 | -0.238 | 0.812 | 0.097 | 6.214 |
| Sex:F (ref=M) | 0.281 | 1.053 | -1.205 | 0.228 | 0.036 | 2.215 |
| Ferritin (<48 ng/mL)*Females | 1.042 | 1.177 | 0.035 | 0.972 | 0.104 | 10.471 |
| Ferritin (48-96 ng/mL) *Females | 14.593 | 1.139 | 2.354 | **0.019** | 1.567 | 135.935 |
| Ferritin (97-168 ng/mL) *Females | 171.066 | 2.501 | 2.056 | **0.040** | 1.270 | 23034.375 |
| Ferritin (>169 ng/mL) *Females | 3.073 | 1.247 | 0.900 | 0.368 | 0.266 | 35.427 |
| Former Smoker (ref= Never Smokers) | 1.319 | 0.392 | 0.706 | 0.480 | 0.612 | 2.844 |
| Current Smoker (ref= Never Smokers) | 4.696 | 0.282 | 5.487 | **<0.001** | 2.703 | 8.160 |
| Prediabetes (ref=No diabetes) | 1.570 | 0.257 | 1.754 | 0.080 | 0.948 | 2.600 |
| Diabetes Mellitus (ref=No diabetes) | 1.337 | 0.403 | 0.720 | 0.472 | 0.607 | 2.945 |
| Non-Hispanic Black (ref: Non-Hispanic White) | 1.296 | 0.398 | 0.651 | 0.515 | 0.594 | 2.829 |
| Mexican-American (ref: Non-Hispanic White) | 1.281 | 0.311 | 0.797 | 0.425 | 0.697 | 2.355 |
| Other Ethnicities (ref: Non-Hispanic White) | 0.251 | 0.740 | -1.865 | 0.062 | 0.059 | 1.072 |
| CRP | 1.810 | 0.259 | 2.293 | 0.022 | 1.090 | 3.004 |
| Age | 1.095 | 0.013 | 7.241 | **<0.001** | 1.068 | 1.122 |
| Creatinine | 1.009 | 0.006 | 1.448 | 0.148 | 0.997 | 1.021 |
| ACR | 0.058 | 1.987 | -1.430 | 0.153 | 0.001 | 2.868 |
| Triacylglycerols | 0.997 | 0.068 | -0.039 | 0.969 | 0.873 | 1.140 |
| BMI | 1.078 | 0.162 | 0.463 | 0.644 | 0.785 | 1.480 |
| HDL | 0.751 | 0.345 | -0.831 | 0.406 | 0.382 | 1.476 |
| Hemoglobin | 0.897 | 0.190 | -0.574 | 0.566 | 0.618 | 1.301 |
| Antihyperlipidemic medications (ref=No) | 1.318 | 0.546 | 0.505 | 0.613 | 0.452 | 3.841 |
| AST | 1.005 | 0.013 | 0.438 | 0.662 | 0.981 | 1.030 |
| ALT | 0.988 | 0.018 | -0.647 | 0.518 | 0.953 | 1.024 |

## Discussion

Our study provides a comparison between ML and regression analyses to demonstrate the association between iron biomarkers in the context of PAD. Taken together, our findings from both analyses show that serum iron biomarkers are associated with PAD, with ferritin demonstrating a consistent association specifically in females with the two methodologies applied.

The positive association for higher serum ferritin among females with PAD strengthens previous observations on the NHANES 1999-2001 cohort^33^. When this observation is compared to other related disease contexts (e.g. carotid/coronary atherosclerosis/cardiovascular disease), ferritin’s positive association agrees with some studies^11,15,16^, but not others^34–36^. Our findings also agree with The Iron and Atherosclerosis Study (FeAST; https://clinicaltrials.gov, NCT00032357), a major trial that investigated the benefits of iron reduction on the clinical outcomes in a cohort of individuals with PAD^37^. While the initial analysis of the FeAST trial did not report a favorable outcome following phlebotomy^37^, the reanalysis of the results adjusting for the effects of age^38^ and smoking^39^ have suggested that iron reduction (with ferritin and TSAT as endpoints) could be of significant benefit for younger individuals and smokers. However, the FeAST trial did not contain sufficient participants to study sex-specific differences. Our study also advances current knowledge. We extend the analyses by Menke et al^33^, by including additional participants from the NHANES 2003-2004 cycle in the analysis. In contrast to the FeAST trial^37^, we have included participants with prediabetes, adjusted more stringently for inflammation [excluded samples with features of inflammation (ferritin>400 ng/mL or CRP>3 mg/dL) and used CRP as a covariate] and increased statistical power to study sex-specific differences.

If the high ferritin values observed in our study were solely interpreted as an indicator of body iron stores, the data suggest that there is an increased risk of PAD at the upper end of, what is clinically accepted as “physiological range”. However, ferritin is also an acute-phase protein and inflammation influences most iron parameters. During inflammatory states, iron export via ferroportin from macrophages and duodenal enterocytes is reduced by hepcidin-controlled^40^ and transcriptional mechanisms^41,42^. Consequently, macrophages become iron loaded contributing to elevated circulating ferritin levels and hypoferremia (low iron levels in the plasma)^43^. Therefore, an alternative interpretation of our findings is that elevated ferritin may be an epiphenomenon secondary to inflammation. In addition, the presence of microinflammation in tissues cannot be completely excluded from the available data.

To summarize, our study adds to the body of evidence suggesting sex-specific differences in PAD^44^ and highlights a possible role of iron (directly or indirectly) in this relationship. We have partly controlled for hormonal influences by excluding NHANES participants who are currently pregnant, menstruating, lactating, or on hormonal contraceptives, but the hormone levels were not available to be included as covariates. It is further possible that additional sex-specific mechanisms are in play.

In both cohorts analyzed, we observed a similar prevalence of PAD (∼6%) associated with well-known risk factors (old age, diabetes, and hypertension). Markers related to metabolic and vascular health, such as blood pressure, plasma glucose levels and CRP, were elevated among PAD participants in both cohorts. Additionally, BMI was higher in PAD participants compared to non-PAD participants in HEIST-DiC, while NHANES shows more consistent BMI values across groups, with a minor increase only in females with PAD. It is important to note that the cohorts analyzed differed in that the HEIST-DiC cohort consisted of mainly European individuals with diabetes and prediabetes from a hospital-based study, while NHANES is a survey cohort, representative of the US population, and therefore contains a mix of ethnicities.

The strength of our study lies in the analyses of these two independent cohorts by orthogonal approaches, allowing for a generalization of the findings, an increased robustness of the data, and a reduction of cohort-specific biases. The examination of multiple iron biomarkers, including serum iron, ferritin, TSAT, and TIBC provides a comprehensive analysis of the relationship between iron and clinically apparent PAD. To the best of our knowledge, this is the first study to evaluate the relationship between iron biomarkers and PAD in prediabetes and diabetes. Our approach also underscores the complementary nature of ML and regression analyses: while ML excels at predictive accuracy and feature selection, regression analyses offer insights into interaction effects and the nuanced relationship between predictors and outcomes. Therefore, integrating both methodologies can leverage the strengths of each approach to provide a comprehensive understanding of complex disease dynamics. Limitations include the cross-sectional nature of the study which limits causal inferences and precludes the assessment of temporal relationships. Further, we were unable to control for all possible confounding factors, such as genetic predisposition, lifestyle, medications, or other comorbidities. The study described here also did not investigate potential mechanisms underlying the observed associations.

## Conclusion

Overall, our data show that elevated ferritin within the physiological range is associated with clinically apparent PAD in females but not males. Further studies are necessary to confirm whether the observed associations are indeed causal and to explore the potential mechanisms underlying this relationship. Our findings need to be considered explorative to encourage the investigation of potential biological pathways underlying the sex-specific differences in iron’s influence on atherosclerosis.

## Supporting information

STROBE Checklist

## Data Availability

The data from this study can be explored here: https://anandr.shinyapps.io/NHANES-HEISTDiC/. The HEIST-DiC dataset analyzed in the current study is available from the corresponding authors on reasonable request. The NHANES dataset is publicly available. Codes used for data analysis and visualization are accessible here (https://github.com/griffindoc/pad)

## Ethics statement

The ethics committees of the University of Heidelberg approved this study (Decision No. 204/2004, 400/2010, and S-383/2016) according to the Declaration of Helsinki. All participants entered the study according to the guidelines of the local ethics committees following written informed consent to participate.

## Author contributions

Anand Ruban Agarvas: Conceptualization, Methodology, Software, Formal analysis, Investigation, Data curation, Writing- Original draft, Visualization

Stefan Kopf: Investigation, Resources, Writing - Review & Editing

Tiago J.S Lopes: Formal analysis, Investigation, Writing- Original draft, Visualization

Paul Thalmann: Validation, Writing - Review & Editing

José Manuel Fernández-Real: Methodology, Writing - Review & Editing, Supervision

Peter Nawroth: Project administration, Funding acquisition, Supervision, Writing - Review & Editing

Martina U. Muckenthaler: Conceptualization, Methodology, Supervision, Writing - Review & Editing, Project administration, Funding acquisition

## Data and Code Availability Statement

The data from this study can be explored here: https://anandr.shinyapps.io/NHANES_HEISTDiC/. The HEIST-DiC dataset analyzed in the current study is available from the corresponding authors on reasonable request. The NHANES dataset is publicly available. Codes used for data analysis and visualization are accessible here (https://github.com/griffindoc/pad)

## Supplementary files

***Supplementary Table 1. Data fields and variables from NHANES 1999-2004.***

The table shows the variables and the corresponding codes used for accessing data from the NHANES. The hyperlinks to the procedure manuals and datafiles are also provided in the table.

**Supplementary Table 2. Multivariable regression analysis of iron biomarkers and PAD in the NHANES Cohort**

We conducted multivariable regression analyses by flexibly modelling the probability of PAD’s presence with iron biomarkers (iron or TSAT) by spline regression (degree of freedom=4). The following variables were included aas covariates: age, CRP, HDLc, triacylglycerols, HbA1c, ACR, BMI, ethnicity (non-Hispanic whites as the reference), smoking (non-smokers as reference), the presence of diabetes, AST, ALT, Hb, creatinine, antihyperlipidemic medications; the former seven variables as continuous and the rest as categorical). All continuous variables were mean-centered for the analysis. In addition, sample weights were included in the analyses

BMI: Body Mass Index, HDL: High-Density Lipoprotein, CRP: High sensitivity C-reactive protein, AST: Aspartate Aminotransferase, ALT: Alanine Aminotransferase; ACR: Albumin-creatinine ratio, OR: Odds Ratio, 95% CI: 95% Confidence Interval, SE: Standard Error

***Supplementary Figure 1. Feature importance of xGBoost classification***

(A-B) We used the XGBoost classifier and its output to quantify the importance of the features to the prediction of PAD occurrence. In both panels, higher values indicate a higher contribution of a given feature, and iron-related markers are color coded, and appear among the top features in terms of relevance to the prediction of this condition.

