## Supplemental Table 1 for "Iron biomarkers predict peripheral artery disease in females: a cross-sectional analysis of HEIST-DiC and NHANES participants"

| **Supplementary Table 1.** | **NHANES 1999-2000** | | | **NHANES 2001-2002** | | | **NHANES 2003-2004** | | |
| --- | --- | --- | --- | --- | --- | --- | --- | --- | --- |
| **Variable name** | **Variable Code** | **Procedure manuals** | **Datafile** | **Variable Code** | **Procedure manuals** | **Datafile** | **Variable Code** | **Procedure manuals** | **Datafile** |
| **Albumin (g/L)** | LBDSALSI | [Link](https://wwwn.cdc.gov/nchs/data/nhanes/1999-2000/labmethods/lab18_met_biochemistry_profile.pdf) | [Link](https://wwwn.cdc.gov/Nchs/Nhanes/1999-2000/LAB18.XPT) | LBDSALSI | [Link](https://wwwn.cdc.gov/nchs/data/nhanes/2001-2002/labmethods/l40_b_met_albumin.pdf) | [Link](https://wwwn.cdc.gov/Nchs/Nhanes/2001-2002/L40_B.XPT) | LBDSALSI | [Link](https://wwwn.cdc.gov/nchs/data/nhanes/2003-2004/labmethods/l40_c_met_albumin.pdf) | [Link](https://wwwn.cdc.gov/Nchs/Nhanes/2003-2004/L40_C.XPT) |
| **Alanine transaminase (U/L)** | LBXSATSI |  |  | LBXSATSI | [Link](https://wwwn.cdc.gov/nchs/data/nhanes/2001-2002/labmethods/l40_b_met_alanine_amino_transferase.pdf) |  | LBXSATSI | [Link](https://wwwn.cdc.gov/nchs/data/nhanes/2003-2004/labmethods/l40_c_met_alanine_amino_transferase.pdf) |  |
| **Aspartate transaminase (U/L)** | LBXSASSI |  |  | LBXSASSI | [Link](https://wwwn.cdc.gov/nchs/data/nhanes/2001-2002/labmethods/l40_b_met_aspartate_aminotransferase.pdf) |  | LBXSASSI | [Link](https://wwwn.cdc.gov/nchs/data/nhanes/2003-2004/labmethods/l40_c_met_aspartate_aminotransferase.pdf) |  |
| **Blood urea nitrogen (mmol/L)** | LBDSBUSI |  |  | LBDSBUSI | [Link](https://wwwn.cdc.gov/nchs/data/nhanes/2001-2002/labmethods/l40_b_met_blood_urea_nitrogen.pdf) |  | LBDSBUSI | [Link](https://wwwn.cdc.gov/nchs/data/nhanes/2003-2004/labmethods/l40_c_met_blood_urea_nitrogen.pdf) |  |
| **Cholesterol, total (mmol/L)** | LBDSCHSI |  |  | LBDSCHSI | [Link](https://wwwn.cdc.gov/nchs/data/nhanes/2001-2002/labmethods/l40_b_met_cholesterol.pdf) |  | LBDSCHSI | [Link](https://wwwn.cdc.gov/nchs/data/nhanes/2003-2004/labmethods/l13_c_met_lipids.pdf) | [Link](https://wwwn.cdc.gov/Nchs/Nhanes/2003-2004/L13_C.XPT) |
| **Triglycerides (mmol/L)** | LBDSTRSI |  |  | LBDSTRSI | [Link](https://wwwn.cdc.gov/nchs/data/nhanes/2001-2002/labmethods/l40_b_met_triglycerides.pdf) |  | LBDSTRSI |  | [Link](https://wwwn.cdc.gov/Nchs/Nhanes/2003-2004/L13AM_C.XPT) |
| **Creatinine (μmol/L)** | LBDSCRSI |  |  | LBDSCRSI | [Link](https://wwwn.cdc.gov/nchs/data/nhanes/2001-2002/labmethods/l40_b_met_creatinine.pdf) |  | LBDSCRSI | [Link](https://wwwn.cdc.gov/nchs/data/nhanes/2003-2004/labmethods/l16_c_met_creatinine.pdf) | [Link](https://wwwn.cdc.gov/Nchs/Nhanes/2003-2004/L40_C.XPT) |
| **Iron (μg/dL)** | LBDIRNSI | [Link](https://wwwn.cdc.gov/nchs/data/nhanes/1999-2000/labmethods/lab06_met_-fe_tibc.pdf) | [Link](https://wwwn.cdc.gov/Nchs/Nhanes/1999-2000/LAB06.XPT) | LBDIRNSI | [Link: Method 1](https://wwwn.cdc.gov/nchs/data/nhanes/2001-2002/labmethods/l40fe_b_met_iron_beckman_synchron.pdf)  [Link: method 2](https://wwwn.cdc.gov/nchs/data/nhanes/2001-2002/labmethods/l40fe_b_met_iron_beckman_synchron.pdf) | [Link](https://wwwn.cdc.gov/Nchs/Nhanes/2001-2002/L40FE_B.XPT) | LBDIRNSI | [Link: Method 1](https://wwwn.cdc.gov/nchs/data/nhanes/2003-2004/labmethods/l40fe_c_met.pdf)  [Link: method 2](https://wwwn.cdc.gov/nchs/data/nhanes/2003-2004/labmethods/l40_c_met_smac_iron.pdf) | [Link](https://wwwn.cdc.gov/Nchs/Nhanes/2003-2004/L40FE_C.XPT) |
| **Transferrin saturation(%)** | LBXPCT | [Link](https://wwwn.cdc.gov/Nchs/Nhanes/1999-2000/LAB06.htm#Analytic_Notes) |  | LBDPCT |  |  | LBDPCT |  |  |
| **Ferritin (μg/L)** | LBDFERSI | [Link](https://wwwn.cdc.gov/nchs/data/nhanes/1999-2000/labmethods/lab06_met_ferritin.pdf) |  | LBDFERSI | [Link](https://wwwn.cdc.gov/nchs/data/nhanes/2001-2002/labmethods/l06_b_met_ferritin.pdf) | [Link](https://wwwn.cdc.gov/Nchs/Nhanes/2001-2002/L06_2_B.XPT) | LBDFERSI | [Link: Method 1](https://wwwn.cdc.gov/nchs/data/nhanes/2003-2004/labmethods/l06tfr_c_met_ferritin_biorad.pdf)  [Link: method 2](https://wwwn.cdc.gov/nchs/data/nhanes/2003-2004/labmethods/l06tfr_c_met_ferritin_hitachi.pdf) | [Link](https://wwwn.cdc.gov/Nchs/Nhanes/2003-2004/L06TFR_C.XPT) |
| **TIBC (μmol/L)** | LBDTIBSI | [Link](https://wwwn.cdc.gov/nchs/data/nhanes/1999-2000/labmethods/lab06_met_-fe_tibc.pdf) |  | LBDTIBSI | [Link: Method 1](https://wwwn.cdc.gov/nchs/data/nhanes/2001-2002/labmethods/l40fe_b_met_iron_beckman_synchron.pdf)  [Link: method 2](https://wwwn.cdc.gov/nchs/data/nhanes/2001-2002/labmethods/l40fe_b_met_iron_beckman_synchron.pdf) | [Link](https://wwwn.cdc.gov/Nchs/Nhanes/2001-2002/L40FE_B.XPT) | LBDTIBSI | [Link: Method 1](https://wwwn.cdc.gov/nchs/data/nhanes/2003-2004/labmethods/l40fe_c_met.pdf)  [Link: method 2](https://wwwn.cdc.gov/nchs/data/nhanes/2003-2004/labmethods/l40_c_met_smac_iron.pdf) | [Link](https://wwwn.cdc.gov/Nchs/Nhanes/2003-2004/L40FE_C.XPT) |
| **Plasma glucose: SI (mmol/L)** | LBXGLUSI | [Link](https://wwwn.cdc.gov/nchs/data/nhanes/1999-2000/labmethods/lab10am_met_plasma_glucose.pdf) | [Link](https://wwwn.cdc.gov/Nchs/Nhanes/1999-2000/LAB10AM.XPT) | LB2GLUSI | [Link](https://wwwn.cdc.gov/nchs/data/nhanes/2001-2002/labmethods/l10am_b_met_glucose.pdf) | [Link](https://wwwn.cdc.gov/Nchs/Nhanes/2001-2002/L10_2_B.XPT) | LBDGLUSI | [Link](https://wwwn.cdc.gov/nchs/data/nhanes/2003-2004/labmethods/l10am_c_met_glucose.pdf) | [Link](https://wwwn.cdc.gov/Nchs/Nhanes/2003-2004/L10AM_C.XPT) |
| **Insulin: SI (pmol/L)** | LBXINSI | [Link](https://wwwn.cdc.gov/nchs/data/nhanes/1999-2000/labmethods/lab10am_met_insulin.pdf) |  | LB2INSI | [Link](https://wwwn.cdc.gov/nchs/data/nhanes/2001-2002/labmethods/l10am_b_met_insulin.pdf) |  | LBDINSI | [Link](https://wwwn.cdc.gov/nchs/data/nhanes/2003-2004/labmethods/l10am_c_met_insulin.pdf) |  |
| **HDL-Cholesterol (mmol/L)** | LBDHDLSI | [Link](https://wwwn.cdc.gov/nchs/data/nhanes/1999-2000/labmethods/lab13_met_lipids.pdf) | [Link](https://wwwn.cdc.gov/Nchs/Nhanes/1999-2000/LAB13.XPT) | LBDHDLSI | [Link](https://wwwn.cdc.gov/nchs/data/nhanes/2001-2002/labmethods/l13_b_met_lipids.pdf) | [Link](https://wwwn.cdc.gov/Nchs/Nhanes/2001-2002/L13_B.XPT) | LBDHDDSI | [Link](https://wwwn.cdc.gov/nchs/data/nhanes/2003-2004/labmethods/l13_c_met_lipids.pdf) | [Link](https://wwwn.cdc.gov/Nchs/Nhanes/2003-2004/L13_C.XPT) |
| **LDL-cholesterol (mmol/L)** | LBDLDLSI |  |  | LBDLDLSI |  |  | LBDLDLSI |  | [Link](https://wwwn.cdc.gov/Nchs/Nhanes/2003-2004/L13AM_C.XPT) |
| **Gender** | RIAGENDR | - | [Link](https://wwwn.cdc.gov/Nchs/Nhanes/1999-2000/DEMO.XPT) | RIAGENDR | - | [Link](https://wwwn.cdc.gov/Nchs/Nhanes/2001-2002/DEMO_B.XPT) | RIAGENDR | [Link](https://wwwn.cdc.gov/Nchs/Nhanes/2003-2004/DEMO_C.htm) | [Link](https://wwwn.cdc.gov/Nchs/Nhanes/2003-2004/DEMO_C.XPT) |
| **Age at Screening** | RIDAGEYR |  |  | RIDAGEYR |  |  | RIDAGEYR |  |  |
| **Race and Ethnicity** | RIDRETH2 |  |  | RIDRETH2 |  |  | RIDRETH2 |  |  |
| **Masked Variance Unit** | SDMVPSU |  |  | SDMVPSU |  |  | SDMVPSU |  |  |
| **Masked Variance Pseudo-Stratum** | SDMVSTRA |  |  | SDMVSTRA |  |  | SDMVSTRA |  |  |
| **Doctor told you have diabetes** | DIQ010 | [Link](https://wwwn.cdc.gov/Nchs/Nhanes/1999-2000/DIQ.htm) | [Link](https://wwwn.cdc.gov/Nchs/Nhanes/1999-2000/DIQ.XPT) | DIQ010 | [Link](https://wwwn.cdc.gov/Nchs/Nhanes/2001-2002/DIQ_B.htm) | [Link](https://wwwn.cdc.gov/Nchs/Nhanes/2001-2002/DIQ_B.XPT) | DIQ010 | [Link](https://wwwn.cdc.gov/Nchs/Nhanes/2003-2004/DIQ_C.htm) | [Link](https://wwwn.cdc.gov/Nchs/Nhanes/2003-2004/DIQ_C.XPT) |
| **Taking insulin now** | DIQ050 |  |  | DIQ050 |  |  | DIQ050 |  |  |
| **Take diabetic pills to lower blood sugar** | DIQ070 |  |  | DIQ070 |  |  | DIQ070 |  |  |
| **Left Ankle Brachial Blood Pressure Index** | LEXLABPI | [Link](https://wwwn.cdc.gov/nchs/data/nhanes/1999-2000/manuals/le.pdf) | [Link](https://wwwn.cdc.gov/Nchs/Nhanes/1999-2000/LEXABPI.XPT) | LEXLABPI | [Link](https://wwwn.cdc.gov/nchs/data/nhanes/1999-2000/manuals/le.pdf) | [Link](https://wwwn.cdc.gov/Nchs/Nhanes/2001-2002/LEXAB_B.XPT) | LEXLABPI | [Link](https://wwwn.cdc.gov/nchs/data/nhanes/2003-2004/manuals/LE.pdf) | [Link](https://wwwn.cdc.gov/Nchs/Nhanes/2003-2004/LEXAB_C.XPT) |
| **Right Ankle Brachial Blood Pressure Index** | LEXRABPI |  |  | LEXRABPI |  |  | LEXRABPI |  |  |
| **Do you now smoke cigarettes** | SMQ040 | [Link](https://wwwn.cdc.gov/Nchs/Nhanes/1999-2000/SMQ.htm) | [Link](https://wwwn.cdc.gov/Nchs/Nhanes/1999-2000/SMQ.XPT) | SMQ040 | [Link](https://wwwn.cdc.gov/Nchs/Nhanes/2001-2002/SMQ_B.htm) | [Link](https://wwwn.cdc.gov/Nchs/Nhanes/2001-2002/SMQ_B.XPT) | SMQ040 | [Link](https://wwwn.cdc.gov/Nchs/Nhanes/2003-2004/SMQ_C.htm) | [Link](https://wwwn.cdc.gov/Nchs/Nhanes/2003-2004/SMQ_C.XPT) |
| **Smoked at least 100 cigarettes in life** | SMQ020 |  |  | SMQ020 |  |  | SMQ020 |  |  |
| **Told had high blood pressure - 2+ times** | BPQ030 | [Link](https://wwwn.cdc.gov/Nchs/Nhanes/1999-2000/BPQ.htm) | [Link](https://wwwn.cdc.gov/Nchs/Nhanes/1999-2000/BPQ.XPT) | BPQ030 | [Link](https://wwwn.cdc.gov/Nchs/Nhanes/2001-2002/BPQ_B.htm) | [Link](https://wwwn.cdc.gov/Nchs/Nhanes/2001-2002/BPQ_B.XPT) | BPQ030 | [Link](https://wwwn.cdc.gov/Nchs/Nhanes/2003-2004/BPQ_C.htm) | [Link](https://wwwn.cdc.gov/Nchs/Nhanes/2003-2004/BPQ_C.XPT) |
| **Taking prescription for hypertension** | BPQ040A |  |  | BPQ040A |  |  | BPQ040A |  |  |
| **C-reactive protein (mg/dL)** | LBXCRP | [Link](https://wwwn.cdc.gov/nchs/data/nhanes/1999-2000/labmethods/lab11_met_c_reactive_protein.pdf) | [Link](https://wwwn.cdc.gov/Nchs/Nhanes/1999-2000/LAB11.XPT) | LB2CRP | [Link](https://wwwn.cdc.gov/nchs/data/nhanes/2001-2002/labmethods/l11_b_met_c_reactive_protein.pdf) | [Link](https://wwwn.cdc.gov/Nchs/Nhanes/2001-2002/L11_B.XPT) | LBXCRP | [Link](https://wwwn.cdc.gov/nchs/data/nhanes/2003-2004/labmethods/l11_c_met_c_reactive_protein.pdf) | [Link](https://wwwn.cdc.gov/Nchs/Nhanes/2003-2004/L11_C.XPT) |
| **Body Mass Index** | BMXBMI | [Link](https://wwwn.cdc.gov/nchs/data/nhanes/1999-2000/manuals/bm.pdf) | [Link](https://wwwn.cdc.gov/Nchs/Nhanes/1999-2000/BMX.XPT) | BMXBMI | [Link](https://wwwn.cdc.gov/nchs/data/nhanes/2001-2002/manuals/body_measures_year_3.pdf) | [Link](https://wwwn.cdc.gov/Nchs/Nhanes/2001-2002/BMX_B.XPT) | BMXBMI | [Link](https://wwwn.cdc.gov/Nchs/Nhanes/2003-2004/BMX_C.XPT) | [Link](https://wwwn.cdc.gov/Nchs/Nhanes/2003-2004/BMX_C.XPT) |
| **Hemoglobin (g/dL)** | LBXHGB | [Link](https://wwwn.cdc.gov/nchs/data/nhanes/1999-2000/labmethods/lab25_met_complete_blood_count.pdf) | [Link](https://wwwn.cdc.gov/Nchs/Nhanes/1999-2000/LAB25.XPT) | LBXHGB | [Link](https://wwwn.cdc.gov/nchs/data/nhanes/2001-2002/labmethods/l25_b_met_complete_blood_count.pdf) | [Link](https://wwwn.cdc.gov/Nchs/Nhanes/2001-2002/L25_B.XPT) | LBXHGB | [Link](https://wwwn.cdc.gov/nchs/data/nhanes/2003-2004/labmethods/l25_c_met_complete_blood_count.pdf) | [Link](https://wwwn.cdc.gov/Nchs/Nhanes/2003-2004/L25_C.XPT) |
| **Platelet count (%) SI** | LBXPLTSI |  |  | LBXPLTSI |  |  | LBXPLTSI |  |  |
| **Taking treatment for anemia/past 3 mos** | MCQ053 | [Link](https://wwwn.cdc.gov/Nchs/Nhanes/1999-2000/MCQ.htm#MCQ053) | [Link](https://wwwn.cdc.gov/Nchs/Nhanes/1999-2000/MCQ.XPT) | MCQ053 | [Link](https://wwwn.cdc.gov/Nchs/Nhanes/2001-2002/MCQ_B.htm) | [Link](https://wwwn.cdc.gov/Nchs/Nhanes/2001-2002/MCQ_B.XPT) | MCQ053 | [Link](https://wwwn.cdc.gov/Nchs/Nhanes/2003-2004/MCQ_C.htm) | [Link](https://wwwn.cdc.gov/Nchs/Nhanes/2003-2004/MCQ_C.XPT) |
| **Ever told you had cancer or malignancy** | MCQ220 | [Link](https://wwwn.cdc.gov/Nchs/Nhanes/1999-2000/MCQ.htm#MCQ220) |  | MCQ220 |  |  | MCQ220 |  |  |
| **Number of times did activity in past 30 days** | PADTIMES | [Link](https://wwwn.cdc.gov/Nchs/Nhanes/1999-2000/PAQIAF.htm#Component_Description) | [Link](https://wwwn.cdc.gov/Nchs/Nhanes/1999-2000/PAQIAF.XPT) | PADTIMES | [Link](https://wwwn.cdc.gov/Nchs/Nhanes/2001-2002/PAQIAF_B.htm) | [Link](https://wwwn.cdc.gov/Nchs/Nhanes/2001-2002/PAQIAF_B.XPT) | PADTIMES | [Link](https://wwwn.cdc.gov/Nchs/Nhanes/2003-2004/PAQIAF_C.htm) | [Link](https://wwwn.cdc.gov/Nchs/Nhanes/2003-2004/PAQIAF_C.XPT) |
| **Metabolic equivalent(MET) intensity level for activity.** | PADMETS |  |  | PADMETS |  |  | PADMETS |  |  |
| **Vigorous activity over past 30 days** | PAD200 | [Link](https://wwwn.cdc.gov/Nchs/Nhanes/1999-2000/PAQ.htm) | [Link](https://wwwn.cdc.gov/Nchs/Nhanes/1999-2000/PAQ.XPT) | PAD200 | [Link](https://wwwn.cdc.gov/Nchs/Nhanes/2001-2002/PAQ_B.htm) | [Link](https://wwwn.cdc.gov/Nchs/Nhanes/2001-2002/PAQ_B.XPT) | PAD200 | [Link](https://wwwn.cdc.gov/Nchs/Nhanes/2003-2004/PAQ_C.htm) | [Link](https://wwwn.cdc.gov/Nchs/Nhanes/2003-2004/PAQ_C.XPT) |
| **Moderate activity over past 30 days** | PAD320 |  |  | PAD320 |  |  | PAD320 |  |  |
| **Donated blood in past 12 months?** | HSQ570 | [Link](https://wwwn.cdc.gov/Nchs/Nhanes/1999-2000/HSQ.htm) | [Link](https://wwwn.cdc.gov/Nchs/Nhanes/1999-2000/HSQ.XPT) | HSD570 | [Link](https://wwwn.cdc.gov/Nchs/Nhanes/2001-2002/HSQ_B.htm) | [Link](https://wwwn.cdc.gov/Nchs/Nhanes/2001-2002/HSQ_B.XPT) | HSQ571 | [Link](https://wwwn.cdc.gov/Nchs/Nhanes/2003-2004/HSQ_C.htm) | [Link](https://wwwn.cdc.gov/Nchs/Nhanes/2003-2004/HSQ_C.XPT) |
| **Are you currently pregnant?** | SEQ060 | [Link](https://wwwn.cdc.gov/Nchs/Nhanes/1999-2000/SEQ.htm#SEQ060) | [Link](https://wwwn.cdc.gov/Nchs/Nhanes/1999-2000/SEQ.XPT) | RHQ141 | [Link](https://wwwn.cdc.gov/Nchs/Nhanes/2001-2002/RHQ_B.htm) | [Link](https://wwwn.cdc.gov/Nchs/Nhanes/2001-2002/RHQ_B.XPT) | RH143 | [Link](https://wwwn.cdc.gov/Nchs/Nhanes/2003-2004/RHQ_C.htm) | [Link](https://wwwn.cdc.gov/Nchs/Nhanes/2003-2004/RHQ_C.XPT) |
| **When did SP have last period?** | RHQ050 | [Link](https://wwwn.cdc.gov/Nchs/Nhanes/1999-2000/RHQ.htm) | [Link](https://wwwn.cdc.gov/Nchs/Nhanes/1999-2000/RHQ.XPT) | RHQ050 |  |  | RHQ051 |  |  |
| **Taking estrogen only pills now?** | RHQ558 |  |  | RHQ558 |  |  | RHQ558 |  |  |
| **Taking progestin only pills now?** | RHQ566 |  |  | RHQ566 |  |  | RHQ566 |  |  |
| **Taking estrogen/progestin now?** | RHQ574 |  |  | RHQ574 |  |  | RHQ574 |  |  |
| **Use estrogen/progestin patch now?** | RHQ600 |  |  | RHQ600 |  |  | RHQ600 |  |  |
| **Now breastfeeding a child?** | RHQ200 |  |  | RHQ200 |  |  | RHQ200 |  |  |
| **Reason not having regular periods** | RHQ040 |  |  | RHQ040 |  |  | RHQ042 |  |  |
| **Creatinine, urine (umol/L)** | URXUCRSI | [Link](https://wwwn.cdc.gov/nchs/data/nhanes/1999-2000/labmethods/lab16_met_creatinine.pdf) | [Link](https://wwwn.cdc.gov/Nchs/Nhanes/1999-2000/LAB16.XPT) | URXUCRSI | [Link](https://wwwn.cdc.gov/nchs/data/nhanes/2001-2002/labmethods/l16_b_met_creatinine.pdf) | [Link](https://wwwn.cdc.gov/Nchs/Nhanes/2001-2002/L16_B.XPT) | URXUCRSI | [Link](https://wwwn.cdc.gov/nchs/data/nhanes/2003-2004/labmethods/l16_c_met_creatinine.pdf) | [Link](https://wwwn.cdc.gov/Nchs/Nhanes/2003-2004/L16_C.XPT) |
| **Albumin, urine (mg/L) SI** | URXUMASI | [Link](https://wwwn.cdc.gov/nchs/data/nhanes/1999-2000/labmethods/lab16_met_urine_albumin.pdf) |  | URXUMASI | [Link](https://wwwn.cdc.gov/nchs/data/nhanes/2001-2002/labmethods/l16_b_met_urine_albumin.pdf) |  | URXUMASI | [Link](https://wwwn.cdc.gov/nchs/data/nhanes/2003-2004/labmethods/l16_c_met_albumin.pdf) |  |
| **Have kidney disease w/ renal dialysis?** | OHQ144 | [Link](https://wwwn.cdc.gov/Nchs/Nhanes/1999-2000/OHXREF.htm) | [Link](https://wwwn.cdc.gov/Nchs/Nhanes/1999-2000/OHXREF.XPT) | OHQ144 | [Link](https://wwwn.cdc.gov/Nchs/Nhanes/2001-2002/OHXREF_B.htm) | [Link](https://wwwn.cdc.gov/Nchs/Nhanes/2001-2002/OHXREF_B.XPT) | OHQ144 | [Link](https://wwwn.cdc.gov/Nchs/Nhanes/2003-2004/OHXREF_C.htm) | [Link](https://wwwn.cdc.gov/Nchs/Nhanes/2003-2004/OHXREF_C.XPT) |
