## Supplemental Table 2 for "Iron biomarkers predict peripheral artery disease in females: a cross-sectional analysis of HEIST-DiC and NHANES participants"

| **Supplementary Table 2** | **Iron vs PAD** | | **TSAT vs PAD** | |
| --- | --- | --- | --- | --- |
|  | **OR [95% CI]** | **P.Value** | **OR [95% CI]** | **P.Value** |
| Intercept | 0 [0-0.032] | **0.005** | 0 [0-0.005] | **<0.001** |
| Sex:F (ref=M) | 1.666 [0.011-253.03] | 0.842 | 1.775 [0.01-322.533] | 0.829 |
| Iron (<11.28 μg/dL) | 0.771 [0.026-22.756] | 0.880 |  |  |
| Iron (11.28-14.85 μg/dL) | 2.785 [0.275-28.228] | 0.386 |  |  |
| Iron (14.86-19.15 μg/dL) | Not estimated |  |  |  |
| Iron (>19.15 μg/dL) | 0.128 [0-102.902] | 0.547 |  |  |
| Iron (<11.28 μg/dL)*Females | 1.237 [0.01-147.893] | 0.931 |  |  |
| Iron (11.28-14.85 μg/dL)*Females | 1.497 [0.039-57.963] | 0.829 |  |  |
| Iron (14.86-19.15 μg/dL)*Females | Not estimated |  |  |  |
| Iron (>19.15 μg/dL) *Females | 0.01 [0-3992.688] | 0.486 |  |  |
| TSAT (<17.7%) |  |  | 1.827 [0.182-18.311] | 0.608 |
| TSAT (17.7-23.5%) |  |  | 25.958 [0.335-2013.133] | 0.142 |
| TSAT (23.6-30.5%) |  |  | Not estimated |  |
| TSAT (>30.5%) |  |  | Not estimated |  |
| TSAT (<17.7%)*Females |  |  | 1.276 [0.012-135.576] | 0.918 |
| TSAT (17.7-23.5%)*Females |  |  | 1.334 [0.001-1683.689] | 0.937 |
| TSAT (23.6-30.5%)*Females |  |  | Not estimated |  |
| TSAT (>30.5%)*Females |  |  | Not estimated |  |
| Prediabetes (ref=No diabetes) | 1.315 [0.839-2.059] | 0.232 | 1.365 [0.886-2.106] | 0.159 |
| Diabetes Mellitus (ref=No diabetes) | 1.27 [0.608-2.655] | 0.524 | 1.301 [0.621-2.729] | 0.486 |
| CRP | 1.631 [0.948-2.806] | 0.077 | 1.738 [1.03-2.935] | **0.038** |
| Age | 1.087 [1.065-1.11] | <0.001 | 1.087 [1.065-1.11] | **<0.001** |
| Creatinine | 1.016 [1.003-1.028] | **0.015** | 1.015 [1.003-1.027] | **0.014** |
| Former Smoker (ref= Never Smokers) | 1.156 [0.571-2.34] | 0.688 | 1.144 [0.555-2.355] | 0.715 |
| Current Smoker (ref= Never Smokers) | 3.418 [1.977-5.91] | **<0.001** | 3.36 [1.901-5.938] | **<0.001** |
| ACR | 0.04 [0.002-0.848] | **0.039** | 0.051 [0.003-0.97] | **0.048** |
| Non-Hispanic Black (ref: Non-Hispanic White) | 1.13 [0.576-2.217] | 0.723 | 1.21 [0.611-2.396] | 0.585 |
| Mexican-American (ref: Non-Hispanic White) | 1.257 [0.715-2.209] | 0.426 | 1.245 [0.713-2.173] | 0.441 |
| Other Ethnicities (ref: Non-Hispanic White) | 0.22 [0.058-0.829] | **0.025** | 0.221 [0.059-0.831] | **0.026** |
| Triacylglycerols | 1.033 [0.892-1.195] | 0.665 | 1.021 [0.876-1.189] | 0.794 |
| BMI | 1.357 [0.905-2.036] | 0.140 | 1.359 [0.906-2.036] | 0.138 |
| HDL | 1.02 [0.481-2.162] | 0.959 | 0.968 [0.473-1.982] | 0.930 |
| Hemoglobin | 1.088 [0.716-1.654] | 0.692 | 1.08 [0.705-1.656] | 0.723 |
| Antihyperlipidemic medications (ref=No) | 1.646 [0.612-4.423] | 0.323 | 1.706 [0.65-4.481] | 0.278 |
| AST | 0.992 [0.965-1.02] | 0.588 | 0.991 [0.964-1.02] | 0.547 |
| ALT | 0.995 [0.959-1.032] | 0.782 | 0.997 [0.96-1.034] | 0.853 |
