## Supplementary figures and images for "Iron biomarkers predict peripheral artery disease in females: a cross-sectional analysis of HEIST-DiC and NHANES participants"

### Supplemental Figure 1

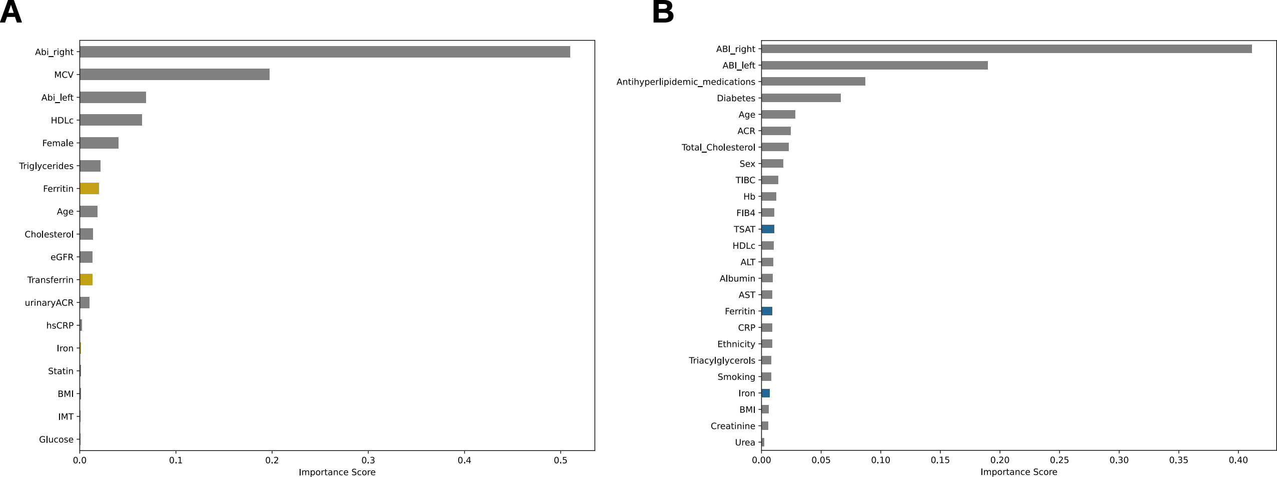
